# Genetic analysis of a malaria outbreak in Laos driven by a selective sweep for *Plasmodium falciparum kelch13* R539T mutants

**DOI:** 10.1101/2022.08.18.22278879

**Authors:** Varanya Wasakul, Areeya Disratthakit, Mayfong Mayxay, Keobouphaphone Chindavongsa, Viengphone Sengsavath, Nguyen Thuy-Nhien, Richard D Pearson, Sonexay Phalivong, Saiamphone Xayvanghang, Sonia Gonçalves, Nicholas P Day, Paul N Newton, Elizabeth A Ashley, Dominic P Kwiatkowski, Arjen M Dondorp, Olivo Miotto

## Abstract

Malaria outbreaks are an important public health concern in endemic regions approaching elimination. Genetic surveillance of malaria parasites can elucidate the population dynamics of an outbreak, and help identify its causes. We investigated the epidemiology of a *Plasmodium falciparum* outbreak in Attapeu Province, Laos, during the 2020-2021 season. An analysis of 249 samples, collected by routine genetic surveillance during the outbreak, revealed a massive loss of genetic diversity in the parasite population, primarily caused by the rapid expansion of a multidrug resistant strain, named LAA1. This strain carried the *kelch13* R539T mutation and expanded clonally, replacing the previously dominant *kelch13* C580Y mutants (KEL1/PLA1) resistant to dihydroartemisinin-piperaquine. Identity by descent (IBD) patterns showed that LAA1 was a recombinant that inherited 60% of its genome from a strain first sampled in Cambodia over a decade ago. A less common outbreak strain (LAA2) carried the *kelch13* C580Y allele, but was distinct from KEL1/PLA1, its genome essentially identical to that of a Cambodian parasite from 2009. A third, low-frequency strain (LAA7) was a recombinant of KEL1/PLA1 with a R539T mutant, the latter providing the *kelch13* variant. These results strongly suggest that the outbreak was driven by a selective sweep, possibly associated with drug-resistant phenotypes of the outbreak strains. The observation that new variants of established multidrug resistant populations can overwhelm previously dominant strains so rapidly has implications for elimination of malaria. Genetic surveillance provides the tools for characterizing outbreaks, and for monitoring the evolution and spread of the populations involved.

## Introduction

The protozoan parasite *Plasmodium falciparum* (*Pf*) is the causative agent of the most severe forms of malaria, causing morbidity and mortality in tropical regions across the globe. In recent years, substantial progress has been made towards reducing the global burden of *Pf* malaria, largely through public health interventions, such as large-scale distribution of mosquito nets that prevent transmission of the parasites via anopheline vectors.^1,2^ At the same time, mortality has been greatly reduced thanks to the availability of highly efficacious artemisinin combination therapies (ACTs) which combine an artemisinin derivative, a fast-acting parasiticidal, with a longer-lasting partner drug to clear surviving parasites from the human host. In the last decade, however, strains resistant to artemisinin have emerged and spread in the Greater Mekong Subregion (GMS), a region of Southeast Asia where transmission is low to moderate.^3-8^ Some of these strains are also resistant to partner drugs,^9-11^ reducing the efficacy of some ACTs and forcing public health authorities to review their choice of frontline therapies. The need to contain the global spread of these resistant strains has prompted intense efforts and major investments to achieve *Pf* elimination from the GMS within the next decade, causing a progressive reduction of falciparum malaria cases across the region. In response to the increased drug pressure, however, the parasite population has continued to evolve and generate new resistant forms which present new challenges to the public health community. As we approach elimination, it has become essential for public health authorities to monitor and characterize epidemiological changes, such as localized malaria outbreaks in areas where progress has been made,^12^ and respond appropriately to combat the resurgence of parasite populations.

Malaria outbreaks are generally thought to be caused by increased transmission, which can be driven by one or more of a number of factors. These include factors related to human behaviour, such as increased travel to forested areas where transmission is more frequent;^13^ or to changing ecology or climate, which affect the size and activity of the mosquito population;^14^ or to the importation of parasites or vectors into areas where elimination had been achieved.^15-17^ A factor that is less frequently considered is that of genetic selection: this may occur when a parasite strain acquires a genetic trait that increases its chances to transmit, and as a result it expands in numbers. Genetic traits may increase transmission rates in various ways-for example, through improved adaptation to local species of vectors, or by increasing the production of gametocytes that can infect mosquitoes. In the GMS, where most infections are symptomatic and thus treated with antimalarials, resistance to these drugs could be a further contributor to transmission increases-for example, if slower parasite clearance rates allow more gametocyte to be produced and passed on to vectors.^6^

Analyses of the genetic patterns characterizing an outbreak population can provide important clues about the factors driving the outbreak. When genetic selection is the driving force, large portions of a genome are co-inherited alongside the selected advantageous mutation, reducing allelic diversity.^18,19^ In parasite populations where self-fertilisation is high, this can lead to rapid expansion of the population under selection, and a collapse of genome-wide variation.^18^ Thus, an outbreak driven by selection may be characterized by a marked reduction in population diversity, decreased heterozygosity and large clonal clusters.^7,20,21^ This is in contrast with outbreaks driven by ecological factors, or by the dynamics of human or vector populations, which would tend to maintain or increase the outcrossing rate, resulting in normal or increased heterozygosity and allelic diversity in the population.^22-24^ In countries nearing elimination, therefore, in-depth analyses of the changes in parasite population structure can be used to discriminate between different outbreak drivers, which can help public health authorities decide on the most effective response-for example, whether to prioritize interventions on forest worker, or investigate frontline treatment efficacy.

The Lao People’s Democratic Republic (Lao PDR, also known as Laos) is a GMS country aiming to eliminate transmission of *Pf* by 2023. Since 2004, the Lao PDR has used artemether-lumefantrine (CoArtem®) as first-line treatment for *Pf*,^25^ which is only endemic in five southernmost provinces, and case numbers have been steadily decreasing over the last few years. However, during the 2020/2021 malaria season, the southern province of Attapeu experienced a sudden increase in the number of *Pf* malaria cases, at a time when other endemic provinces in the country saw a marked reduction.^26^ The GenRe-Mekong project has been conducting routine genetic surveillance of malaria in southern Laos since 2017 in collaboration with the Centre of Malariology, Parasitology, and Entomology (CMPE) of the Lao PDR, and collected 249 samples from cases in Attapeu Province during the outbreak. These samples were genotyped with a broad set of markers,^27^ and their genetic signatures were compared with those of parasites from previous seasons and other geographical locations. Here, we describe the results of the genetic analyses carried out to characterize the strains associated with the Attapeu outbreak, reconstruct the population dynamics of this outbreak, and provide information about the origins of these strains. In spite of operational obstacles presented by the COVID-19 pandemic, the streamlined genotyping approach allowed GenRe-Mekong to return analysis results before the start of the following malaria season, providing a near-real time response to the outbreak, and informing interventions by public health authorities in the Lao PDR and neighbouring countries.

## Results

### The Attapeu outbreak was characterized by a massive drop in genetic diversity

The southern Lao province of Attapeu experienced a rise in the number of cases during 2020-2021, in sharp contrast with a low *P. falciparum* incidence in other endemic parts of the country, including Savannakhet Province where the highest proportion of cases had been seen in previous years. This epidemiological change was reflected by the number of samples collected by GenRe-Mekong (Supplementary Figure 1). To investigate this outbreak, we genotyped all samples collected in this province before lockdowns came into effect due to the COVID-19 pandemic, resulting in a set of 249 *P. falciparum* surveillance samples, from cases presenting at public health facilities between 1 April 2020 and 31 March 2021. These were used in a series of analyses in which they were compared to 2078 samples collected during routine surveillance in Attapeu (n=622) and other provinces of southern Laos since 2017, and 86 samples from Attapeu collected by the TRAC clinical study in 2011-2012.^6^ To support analyses of diversity and genetic distance, we used data from genetic barcodes comprising 101 SNPs selected for their variability and low mutual linkage (see Methods).^27^ As a quality filtering step, we excluded samples with a high degree of genotype missingness (>25%), resulting in a final set of 1787 samples, including 190 collected during the outbreak (Supplementary Table 1).

As a first step towards characterizing the outbreak parasite populations, we compared the diversity of genetic barcodes in this population with that of parasites circulating in Attapeu in previous years, using the average SNP heterozygosity across all barcoding loci as a measure of diversity (see Methods). This showed that diversity in the period 2017-2019 was essentially stable and similar to that observed in the period 2011-2012, with mean heterozygosity in the range [0.39-0.42]. Starting in 2020, however, the levels of diversity collapsed, reaching a level of heterozygosity of 0.12 in 2021 (t=18.7, p<0.001 when comparing Attapeu samples in 2020-2021 vs 2012-2019, Figure 1A). Prior to the outbreak, diversity in other provinces was somewhat lower (range=[0.36-0.38]) than in Attapeu (0.41; p=0.023) but still significantly higher than in the most recent Attapeu samples (F=71.3, p<0.001, Supplementary Figure 2). Combining these perspectives, we can conclude that the outbreak was characterized by a massive loss of diversity, out of the ordinary both for Attapeu and for Laos in general. In this context, it is worth noting that Savannakhet Province had experienced seasonal case number rises in the three previous seasons (Supplementary Figure 1), but these epidemics were not accompanied by massive diversity reduction, suggesting that their drivers were of a different nature from those of the Attapeu outbreak.

**Figure 1.**
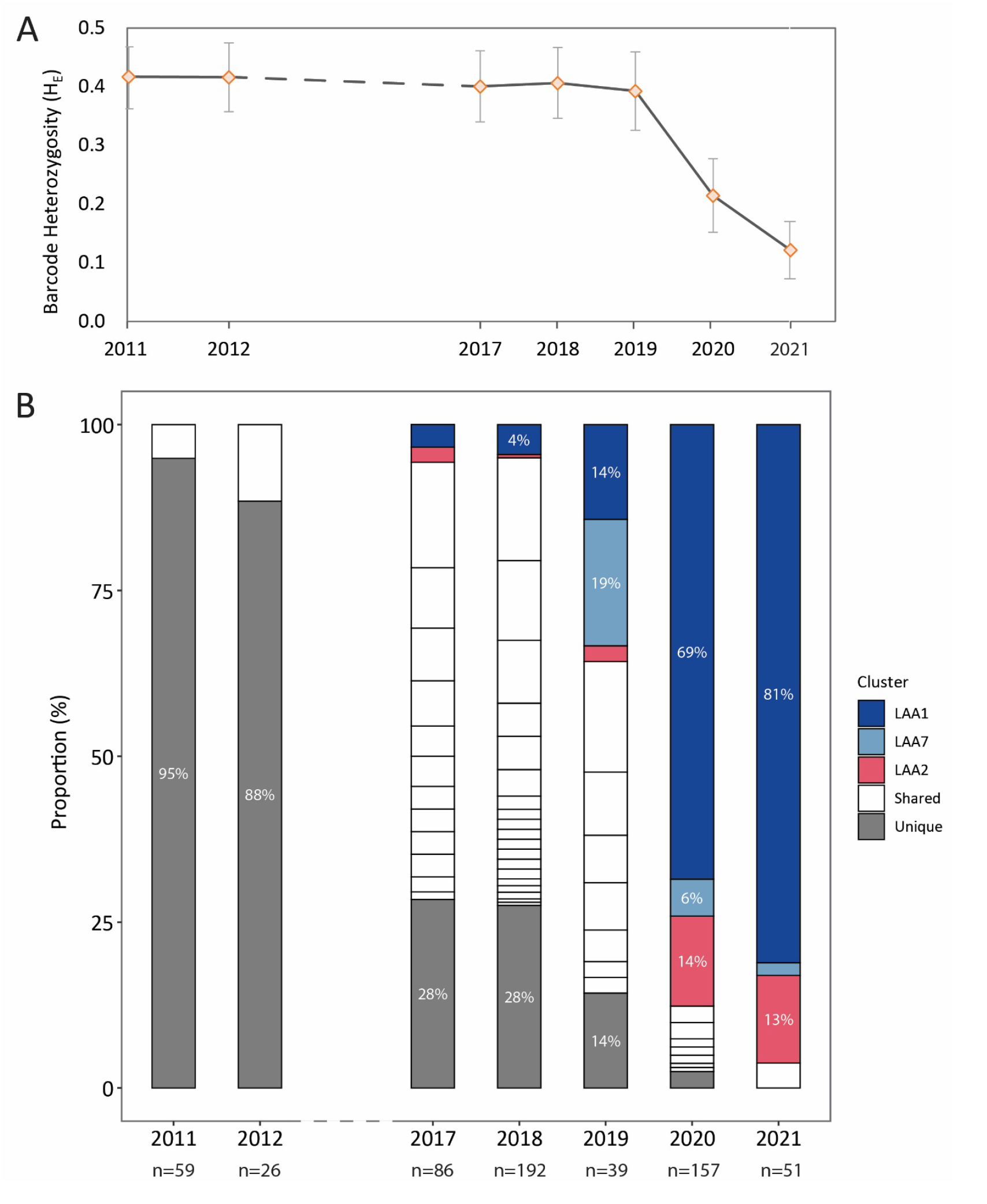
Temporal changes in the population diversity in Attapeu province. (A) Changes in population diversity over time. Mean SNP heterozygosity across all genetic barcode loci is estimated as a measure of diversity for each year. The dashed line indicates a 5-year gap where data is not available. Error bars show standard deviation. (B) Increasing prevalence of clusters with highly related parasites (≥ 95% identity), and decreasing prevalence of parasites with unique barcodes (not belonging to a cluster). Bar segments represent the clusters identified each year, their size being proportional to the number of cluster members; parasites with unique barcodes are grouped in a gray segment at the bottom of the bar. Clusters that dominates the population during the 2020-2021 outbreak (LAA1, LAA2 and LAA7) are shown in colour. The total number of samples (n) in each year is shown below the bars.

### The diversity collapse was driven by a clonal expansion of *kelch13* R539T mutants

Since such marked drop in diversity is likely to be underpinned by one or more expanding populations, we sought to further clarify the changes in population structure that accompanied the outbreak. To identify populations of highly related parasites, we calculated pairwise genetic distances based on a comparison of genetic barcode, and used these data to cluster together parasites with at least 95% barcode identity (see Methods). This identified 30 clusters of varying sizes (>2 members) within the Attapeu sample set. In 2011-2012, before the regional spread of the multidrug-resistant KEL1/PLA1 parasites that migrated into the southernmost provinces of Laos,^27,28^ the diversity of genetic barcodes was very high, such that at least 88% of parasites did not cluster with any other parasite (Figure 1B). In the period 2017-2018, considerably more population structure emerged, as parasites formed several small clusters, and the proportion of parasites carrying a unique barcode dropped to ∼28%. Several of these low-diversity clusters may represent sub-strains of KEL1/PLA1, as previously described.^28^ However, starting in 2019 and leading into the outbreak period, many of these disappeared, as three clusters (labelled LAA1, LAA2 and LAA7) grew to dominate the population. Some parasites in these clusters appeared to be in circulation before 2019 at low frequency, but subsequently expanded so aggressively as to constitute over 95% of the individuals sampled in 2021. The most rapidly expanding cluster (LAA1) rose to 81% of the population in 2021, and must clearly be responsible for most of the observed diversity reduction. The second largest cluster (LAA2) expanded more gradually, reaching a frequency of 14% in 2021, while the third (LAA7) appears to have shrunk in size as LAA1 expanded. To summarize, the *P. falciparum* population in Attapeu has undergone a progressive increase in population structure, initially likely to have been driven by imported artemisinin-resistant strains. However, in the 2019-2021 period, a very small number of parasites clusters has effectively replaced the previous populations through what amounts to clonal expansions, strongly suggesting that the outbreak is a result of a selective sweep.

To look for possible drivers of genetic selection, we labelled samples according to their genotypes at some key drug resistance markers, and characterized clusters accordingly (Supplementary Table 2). As expected, several of the clusters present in the years 2017-2018 comprised multidrug-resistant parasites possessing the *kelch13* C580Y mutation and the *plasmepsin 2/3* (*pm23*) amplification (markers of resistance to artemisinin and piperaquine, respectively), and therefore likely to be variants of the KEL1/PLA1 strain.^28^ This strain proliferated in several GMS countries that deployed the ACT dihydroartemisinin-piperaquine (DHA-PPQ) as frontline treatment, and is known to have reached the southernmost provinces of Laos, even though DHA-PPQ was not used in this country.^27^ During the outbreak, KEL1/PLA1 rapidly disappeared and was replaced by the three outbreak clusters (Supplementary Figure 3). All three are multidrug resistant strains possessing markers associated with resistance to chloroquine, sulfadoxine, pyrimethamine, and artemisinin (Supplementary Table 2). The LAA2 cluster carried the C580Y mutation, although the absence of the *pm23* amplification suggests it might not be closely related to the KEL1/PLA1 parasites. The remaining two clusters (LAA1 and LAA7) both comprised parasites carrying the R539T *kelch13* mutation, without *pm23* amplification. Therefore, a rapid switch in prevalence of *kelch13* mutations was observed in 2020, with R539T mutants prevailing over those carrying C580Y, and causing the excess of cases (Figure 2A). Consistent with this, we found the R539T mutation to be very strongly associated with the emergence of the outbreak (p < 0.0001, Supplementary Table 3). Since the geographical extent of LAA1 and LAA7 appears limited to Attapeu Province (Supplementary Figure 4), it is likely that these lineages have emerged locally. The *kelch13* R539T mutation was not detected in other endemic provinces of Laos (Figure 2B), but was present at low frequency in Attapeu before the outbreak.

**Figure 2.**
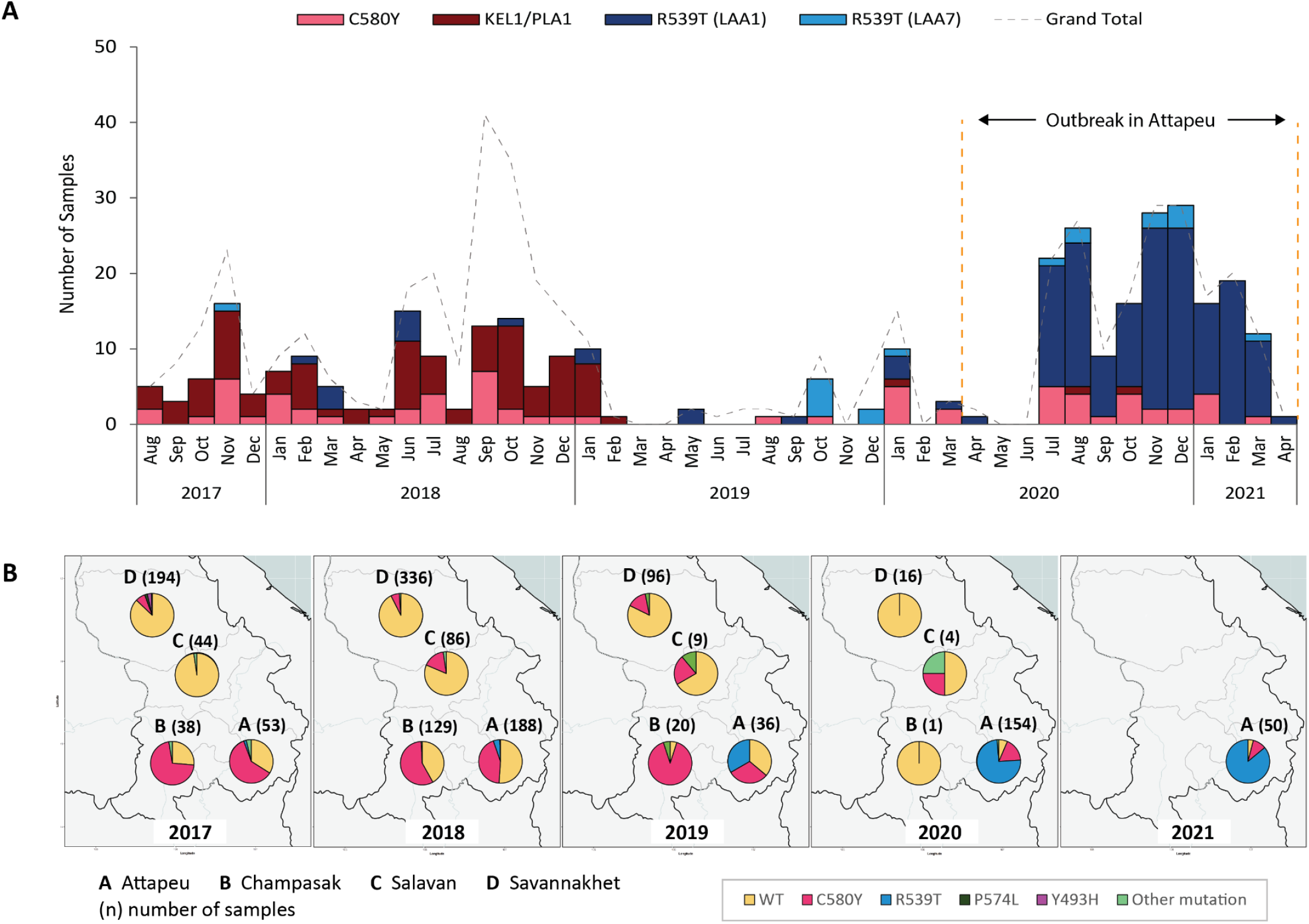
Prevalence of *kelch13* mutations. (A) Temporal distribution of selected *kelch13* mutations in Attapeu province, for each month from August 2017 to April 2021. The bars show the counts for: C580Y mutants without *plasmepsin 2/3* amplification (pink), C580Y mutants with *plasmepsin 2/3* amplification (KEL1/PLA1, dark red), LAA1 R539T mutants (dark blue), and LAA7 R539T mutants (light blue). A dashed line shows the total number of samples collected in the same periods (i.e. including samples with wild-type *kelch13* and others). (B) Spatiotemporal distribution of *kelch13* mutations in four provinces of southern Laos (Attapeu, Champasak, Salavan and Savannakhet). Number of samples in each province are shown in brackets. Sekong province was not included due to its low number of samples.

### The LAA1 outbreak strain was not closely related to other Attapeu clusters

To better understand the relationship between parasite clusters, we visualized genetic distances between pairs of individuals, by constructing a relatedness heatmap (Figure 3). Its most prominent feature is the extremely high degree of similarity among members of the large LAA1 cluster (median pairwise distance within the cluster=0.0), showing very little evidence of recombination between this essentially clonal group and other clusters. At the same time, LAA1 appear to diverge from the rest of Attapeu parasites (median pairwise distance between LAA1 and non-LAA1 parasites=0.42, p<0.001). Although clonality and differentiation are features typical of a founder effect, we note that LAA1 individuals circulated in small numbers prior to the LAA1 expansion, and did not prevail for several seasons, supporting the notion that the expansion is primarily the result of a selective sweep.

**Figure 3.**
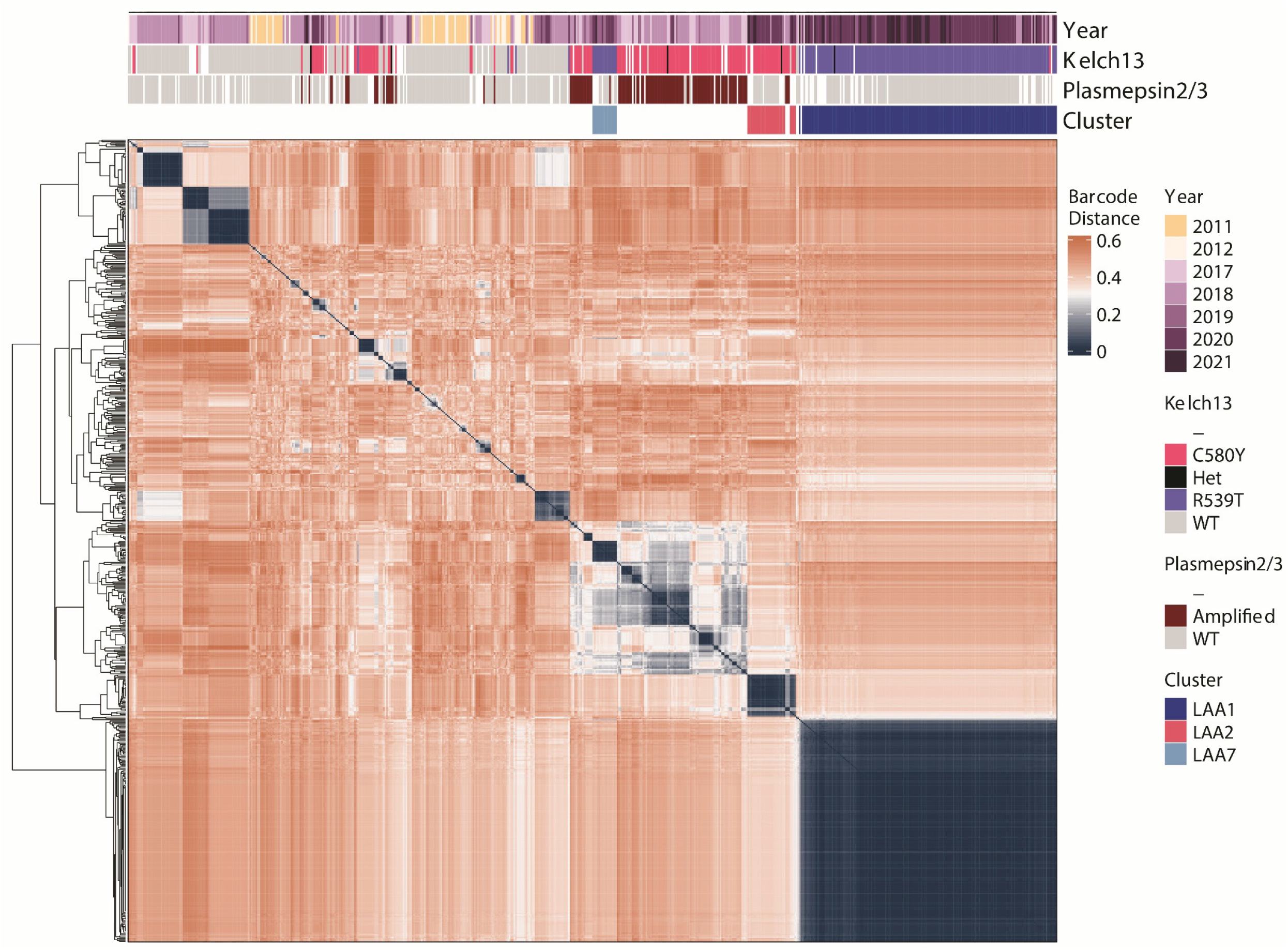
Heatmap of relatedness between populations in Attapeu based on pairwise genetic barcode distances. A pairwise distance matrix was calculated from a set of 610 samples from Attapeu, using 96 quality-filtered barcode SNPs. The arrangement of samples on the heatmap is determined by hierarchical clustering; the dendrogram on the left shows the clustering hierarchy. Colour bars at the top of the heatmap indicate for each sample: the year of sampling; the *kelch13* genotype; the *plasmepsin 2/3* amplification status; and the cluster to which the sample belongs. Heatmap cell colours reflect the pairwise distances between samples: shades of blue represent high pairwise similarity, the darkest blue corresponding to identical genetic barcodes; shades of red signify low barcode similarity, with deepest red representing a distance of 0.6 (i.e. 60% of the barcode alleles differ between a pair of samples).

The characteristics of LAA1 can be contrasted with those of the KEL1/PLA1 clusters, which group together a fragmented set of related clusters in the heatmap, consistent with the presence of differentiated sub-lineages. Surprisingly, LAA7 is also located within this group of clusters, although it carries a R539T allele rather than C580Y, and does not possess the *pm23* amplification-the two main features that characterize KEL1/PLA1. This suggests that LAA7 may have originated from KEL1/PLA1 parasites that, through recombination, have acquired a different *kelch13* variant and lost the *pm23* amplification. The LAA2 outbreak cluster, on the other hand, does carry the *kelch13* C580Y mutation but does not group with the KEL1/PLA1 clusters, which could indicate that it derives from a separate C580Y mutant lineage. Additional analyses using principal coordinates analysis (PCoA) corroborated these observations, showing that LAA1 clearly sets itself apart from other clusters; that LAA2 is distinct from earlier KEL1/PLA1 clusters; and that LAA7 is more closely related to the C580Y parasites than to LAA1 (Supplementary Figure 5).

### The outbreak strains are descendants of early lineages from Cambodia

The above findings raise questions about the origins of the clusters that caused the outbreak, particularly those carrying the *kelch13* R539T mutation. Did these strains emerge as a result of independent mutation events, or are they descendants of long-standing drug resistant strains, previously reported in other parts of the GMS?^20,29,30^ Can recombination events explain the similarities between LAA7 and the KEL1/PLA1 strains? To answer these questions, we reconstructed the ancestry of the three outbreak strains using *identity by descent* (IBD) analysis. This method compares the genomic sequences of pairs of individuals, identifying stretches of the genome likely to have been inherited from a common ancestor, since the identity between the two parasites cannot be statistically explained by chance. For this analysis, we used whole-genome sequence data from parasites from Laos and its neighbouring countries, contributed to the MalariaGEN *P. falciparum* Community Project V6.4, which included a small number of LAA1, LAA2 and LAA7 samples collected by the GenRe-Mekong surveillance project (Supplementary Table 4).

The IBD analysis findings are summarized in Figure 4. As a starting point, we considered some of the oldest *kelch13* mutants available, particularly the early founder populations circulating in western Cambodia over a decade ago. Two of these populations, labelled KH2 and KH3 in previous work,^20^ were subsequently found to harbour C580Y and R539T mutants respectively.^30^ When comparing the LAA1 with one of the R539T mutants (KH3, collected in western Cambodia in 2008), we found 58.8% IBD between the genomes, including a region containing the *kelch13* gene (Supplementary Figure 6). This indicates that the artemisinin resistance mutation carried by LAA1 has not emerged independently in Laos, but was inherited from strains circulating over a decade ago. The segmentation of IBD in large regions of the genome indicates limited linkage breakdown, implying that very few recombination events have occurred during this lengthy time interval. This was confirmed by an analysis of more recent R539T mutants, showing that parasites essentially identical to Cambodian KH3 (KH3-LA, IBD ∼100%) were circulating in Laos in 2017. In other words, this lineage of R539T mutants has circulated and spread across national borders, essentially unchanged, for almost a decade, and was the majority contributor in recombination events that produced LAA1 in Attapeu Province, likely around 2017 when the earliest LAA1 members were sampled. Although we could not identify a single donor for the remaining regions of the LAA1 genome, a *kelch13* wild-type parasite from Champasak Province (LAA1-preWT) exhibited 27.1% IBD with LAA1, including regions distinct from the KH3 contributions, suggesting that the recombination events that produced LAA1 involved local wild-type parasites. In contrast, a KH2 C580Y mutant (KH2A, collected in western Cambodia in 2009) only shared 8.3% IBD with LAA1 ancestry, some of which overlapped with the KH3 contribution (Supplementary Figure 6).

**Figure 4.**
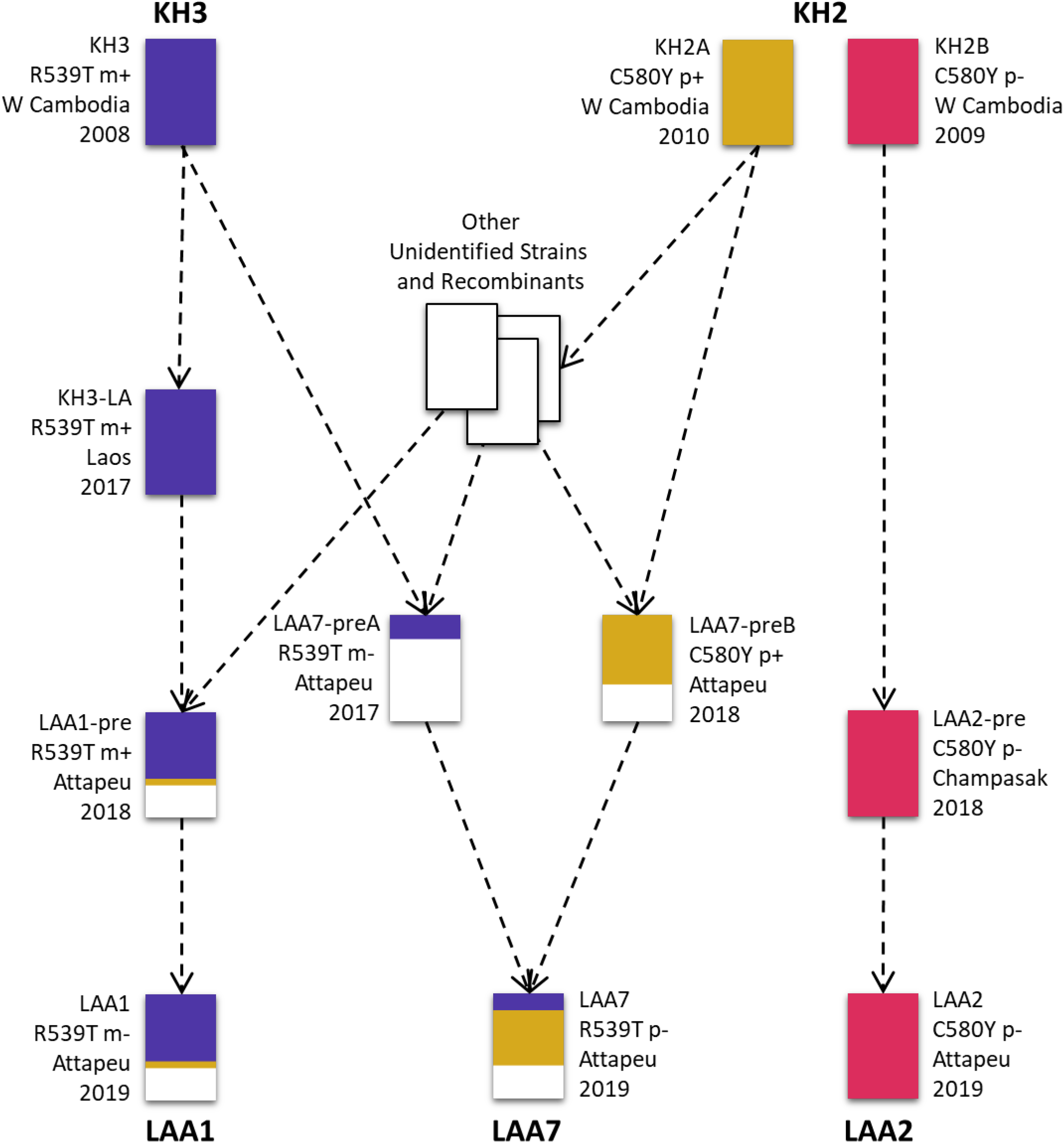
Reconstruction of ancestry of the Attapeu outbreak strains. The diagram shows the relationship between strains analysed for IBD (listed in Supplementary Table 4). Each rectangle represents a sequenced sample: the three samples at the top represent older kelch13 mutants from Cambodia, the three samples at the bottom represent the Attapeu outbreak clusters; the remaining rectangles represent intermediate descendants and other parasite strains. Each rectangle is filled with coloured segments that show the proportion of contribution from the older Cambodian strains; white segments represent contributions from unknown sources. Next to each rectangle, we indicate: the sample label, the *kelch13* mutation and *plasmepsin 2/3* amplification status (p-indicates a single copy, p+ multiple copies), the place and year of sampling. Dashed arrows suggest putative inheritance paths, derived from observing IBD patterns, but may not denote an actual direct descendance of one sample from another (as opposed to both having common ancestry). Furthermore, it should be clear that numerous intermediate ancestry steps are not shown in this highly simplified depiction.

The ancestry of LAA2 was relatively simple to reconstruct: its genome is essentially identical to that of the KH2A parasite (IBD > 99%), suggesting that LAA2 is a recent expansion of a strain that has spread regionally, since 2009 or earlier, probably from western Cambodia, approximately 500 kilometres away from Attapeu. Although these parasites carry the C580Y mutation, they do not possess the *pm23* amplification which characterizes the KEL1/PLA1 lineage. This is confirmed by the lower IBD level (36.9%) observed when comparing LAA2 with an early C580Y mutants carrying the amplification (KH2B, collected in western Cambodia in 2010), indicating that LAA2 derived from a lineage related to but distinct from KEL1/PLA1.

IBD analyses showed that the LAA7 cluster is likely to have originated from the recent recombination of two local strains, one carrying the R539T mutation (LAA7-preA, sampled in Attapeu, 2017) and the other a C580Y mutant (LAA7-preB, Attapeu, 2018), which contributed 49.0% and 45.1% respectively (Supplementary Figure 7). The two donor strains appear to be themselves recombinants with substantial portions of their genome in IBD with the early Cambodian mutants. LAA7-preA shares 23.1% of its genome with KH3, including the region containing the R539T *kelch13* variant, which was subsequently inherited by the LAA7 strain; we were not able to reliably assign other sources to the remainder of the LAA7-preA genome. The LAA7-preB parasites, on the other hand, appear to be a strain of KEL1/PLA1 parasites with a high degree of similarity with earlier Cambodian strains (65.5% IBD with KH2B). This result is consistent with the KEL1/PLA1 lineage differentiation described previously.^28^ The contribution of LAA7-preB has resulted in a high level of IBD between LAA7 and KEL1/PLA1 genomes, explaining the relationship observed in the genetic distance heatmap.

## Discussion

In malaria endemic regions approaching elimination, outbreaks threaten progress and can result in a resurgence of the disease. In this study, we have shown that genetic surveillance offers an informative new perspective on outbreaks, and can shed light on their driving forces. Crucially, it provides insights from the parasite population viewpoint, allowing us to categorize the strains involved, and understand how they have evolved and interacted with each other. This was achieved by studying three key aspects: the genetic diversity at population level, the presence of markers of drug resistance, and the recombination patterns that produced the outbreak strains.

The study of genetic diversity in the Laos outbreak showed that the increase in the number of cases was primarily caused by the clonal expansion of a single multidrug resistant strain, carrying the *kelch13* R539T mutation that confers resistance to artemisinin. This expansion presented some interesting and revealing features. First, it occurred very rapidly, in a single season during which the previously dominant population of KEL1/PLA1 parasites was effectively replaced. KEL1/PLA1 strains had become established in Champasak and Attapeu Provinces, probably following importation from neighbouring countries where DHA-PPQ was the ACT of choice. Our data shows that, although LAA1 parasites were present since 2018, they remained at low frequency while KEL1/PLA1 dominated. This changed in 2019-2020, when the KEL1/PLA1 population in Attapeu collapsed, leaving a vacuum that was rapidly filled by the sudden rise in frequency of LAA1. It is noteworthy that these events coincided with the end of DHA-PPQ usage in neighbouring provinces of northeast Thailand, which perhaps relieved selective pressure on the parasite population.

Second, the expansion was essentially clonal, with very little evidence of recombination with other strains, which is characteristic of a population under strong selective pressure. Had the outbreak been driven by increased transmission due to environmental or human factors, we would have expected a considerably higher level of recombination in Attapeu Province, and more diversity. The very low level of diversity seen in the outbreak was highly unusual, when compared both with other provinces of Laos, and with previous seasons in Attapeu. It does not seem plausible that recombination was limited by a lack of parasite variety, since we observed two additional clonally expanding clusters, and some wild-type parasites, circulating during the outbreak. Such low levels of recombination could occur if LAA1’s selective advantages were conferred by a complex set of mutations, distributed across the genome, that would be disrupted when recombining with parasites with a different genetic background. In this case, parasites resulting from selfing (i.e. from mating with genetically identical individuals) would retain the selective advantage and have a greater chance of survival than other recombinants. This may also explain how the LAA2 outbreak strain has preserved its genome unchanged for at least 12 years since it was first sampled in Cambodia-strongly suggesting that LAA2 possesses a set of genetic variants that is most advantageous when kept whole. As a result of continued selfing, this lineage has retained its competitiveness, without prevailing in any specific place or period of time, and surviving the dominance of other strains, to enjoy a revival after more than a decade, hundreds of kilometres away from its origins. This is by no means a unique case: at the origin of the LAA1 population we have identified practically identical individuals belonging to the KH3 strain, sampled in different countries almost a decade apart. Although we have also found evidence of recombination events involving these strains, IBD patterns suggest that successful recombinants are infrequent, as evidenced by the limited linkage breakdown in the LAA1 and LAA7 genomes.

All evidence points to a selective sweep as the cause of the Attapeu outbreak. What the data cannot tell us at this stage is the exact nature of the selective advantage that contributed to the expansion of LAA1 and other outbreak populations. All factors that increase a parasite’s chance to transmit, such as increased gametocyte production, adaptation to local mosquito populations, and so on, may be drivers of selection. However, given the recent history of malaria in the GMS, antimalarial drug resistance seems the most likely driver of selective sweeps; specifically, one might suspect that LAA1 parasites have reduced sensitivity to lumefantrine, the frontline ACT partner drug in Laos. This can neither be confirmed nor refuted by our data, since there is no widely accepted and validated marker of lumefantrine resistance; most LAA1 parasites even lack the *mdr1* gene amplification, a proposed candidate marker,^31^ although this amplification was a feature of their R539T mutant precursors. Still, it is certainly possible that unknown drug resistance mutations may have been inherited by LAA1 from unknown contributors. We strongly recommend that the efficacy of artemether-lumefantrine be confirmed in Attapeu Province, and it would also be valuable to test the *in vitro* response to individual drugs.

Our analysis of IBD indicates that, to this day, novel artemisinin-resistant lineages are still inheriting *kelch13* haplotypes from early mutants that circulated in western Cambodia. In other words, in spite of the proliferation of artemisinin-resistant parasites, the *kelch13* mutations in circulation in the eastern GMS originate from a surprisingly small number of mutation events,^30^ in contrast with the wider variety of independently emerged mutations observed in Myanmar.^32,33^ It is possible that the small number of founder populations described a decade ago^20^ had already gone through a selection process that had weeded out less fit mutants. An important take-home lesson from the Attapeu outbreak is that these populations and their derivatives have been very resilient over time. Even while KEL1/PLA1 parasites seemed to head for complete domination,^34^ R539T parasites and C580Y parasites without *pm23* amplification have remained in circulation, stealthily waiting for their opportunity to prevail. Such alternation in prevalence is most likely to be triggered by human behaviours affecting selective pressures, such as changes in the choice of frontline treatment, or targeted elimination interventions. Therefore, it remains crucial for genetic surveillance in this region to monitor all *kelch13* mutant varieties, rather than focus on specific variants whose dominance may be temporary. Using whole-genome sequencing data, it may be feasible to put together a systematic catalogue of drug resistant strains and the genetic markers by which they can be tracked and mapped. Here, we have shown that ancestry reconstruction methods, such as IBD inference, can be powerful tools for this type of investigation.

The analyses presented in this paper have demonstrated that routine genetic surveillance, which is relatively straightforward and affordable to implement,^27^ is a powerful tool that can provide deep insight into the causes and dynamics of malaria outbreaks. Using simple 101-SNP barcodes, we identified three different expanding strains and showed that diversity had dropped to abnormal levels, indicative of a selective sweep underpinning the outbreak. By genotyping genetic markers of drug resistance available, we were then able to characterize these populations. Crucially, rapid processing enabled us to communicate our findings with the National Malaria Control Programme before the start of the following season, so that they could plan their response. The subsequent addition of whole-genome sequencing data has enabled a further in-depth study of the origin of the outbreak populations, and their relationship to other well-known resistant strains and to unknown local parasites. We believe there is much value in analysing genomic epidemiology at different levels of resolution, using simpler techniques to provide rapid answers to pressing questions, and more detailed methods to develop a deeper understanding of the underlying phenomena and processes, with which to tackle outbreaks more effectively in the future. We encourage the genomic epidemiology and surveillance communities to continue contributing to shared genomic datasets that will continue to support tackling these and other important public health use cases.

## Methods

### Ethics Statement

This study was conducted as part of the GenRe-Mekong project for the genetic surveillance of malaria in southern Laos, using a variant of the GenRe-Mekong protocol which is approved by the relevant ethics committees in each country where sample collections are carried out; for further details, refer to the GenRe-Mekong manuscript.^27^ For Laos, ethical approvals were obtained from the National Ethics Committee for Health Research (NECHR) of the Health Ministry of the Lao People’s Democratic Republic and the Oxford Tropical Research Ethics Committee (OxTREC).

### Sample and data collection

Between 1 January 2017 and 31 March 2021, a total of 2078 *Plasmodium falciparum* samples were collected at 55 local public health sites in the southern Laos provinces of Savannakhet, Salavan, Sekong, Champasak and Attapeu. Samples were collected from patients of all ages who has been confirmed positive for *P. falciparum* by rapid diagnostic test or blood smear microscopy at partner public health sites (see Supplementary File for a full list of sites and credits to local collaborators). After obtaining consent, three 20 µl dried blood spots (DBS) on filter paper were obtained from each patient by finger prick, dried and stored in a plastic bag with silica gel. The DBS samples, sample manifests and site records were labelled with scannable unique identification tags. The sample collection date and sample-related information were recorded in the sample manifests and site records. Details of sample collection procedure by GenRe-Mekong has been fully described elsewhere.^27^

For ancestry analyses, we used additional whole-genome sequence (WGS) data from parasites from Laos, Cambodia, Thailand and Vietnam included in the public release of the MalariaGEN *P. falciparum* Community Project (PfCP) V6.0,^35^ as well as WGS data from Laos samples contributed by GenRe-Mekong and genotyped in subsequent partner releases. All samples were genotyped using the PfCP 6.0 variant list. Supplementary Table 4 provides information on the WGS samples used in this paper.

### Genotyping and whole-genome sequencing

Genomic DNA was extracted from dried blood spot using high-throughput robotic equipment (Qiagen QIAsymphony) according to manufacturer’s instructions. Parasite DNA was amplified using selective whole genome amplification (sWGA)^36^ before targeted genotyping and whole genome sequencing (WGS). Targeted genotyping was performed by the SpotMalaria platform as previously described.^27^ For each sample, we produced a Genetic Report Card (GRC) detailing calls for a standardized set of variants and haplotypes relevant to drug resistance, as well as a genetic barcode to enable epidemiological analyses. Full details of GRC contents, and how they are derived through targeted genotyping, are given elsewhere.^27^ For samples for which targeted genotyping was not available, GRC variants were called from WGS data using bespoke scripts.

WGS data were obtained using Illumina short-read sequencing technology. Due to lengthier processing times and to operational restrictions during the COVID-19 pandemic, WGS data were not available for samples collected during the later stages of the outbreak. Genotypes were called from WGS reads at 1,042,396 nuclear biallelic single-nucleotide polymorphisms (SNPs) classified as PASS in the PfCP v6.0 variants list (https://www.malariagen.net/data/catalogue-genetic-variation-p-falciparum-v6.0), using the standardised MalariaGEN *Plasmodium falciparum* Community Project genotyping pipeline V6.0.^35^

For a number of samples, we performed copy number estimation for the *plasmepsin2/3* and/or *mdr1* genes by quantitative real-time PCR. The *plasmepsin2/3* amplification was called either by detection of a duplication breakpoint^35^ or by qPCR estimation^37^. The *mdr1* amplification was detected either by qPCR estimation^31^, or from WGS data using a coverage-based method.^35^

### Quality filtering of sample data

As part of the GRC, each sample is assigned a genetic barcode comprising 101 SNP alleles, chosen based on their variability and their power to recapitulate genetic distance.^27^ Before proceeding with epidemiological analyses, we performed a quality filtering step to reduce errors due to missingness in the data (i.e. positions where a genotype could not be called). First, we removed samples with more than 50% missing barcode genotypes. Then, we filtered out barcode SNPs with missing calls in more than 25% of the remaining samples. Lastly, we removed samples with more than 25% missing calls in the remaining barcode SNPs.

### Pairwise genetic distances based on genetic barcodes

To identify clusters of highly related parasites and provide input for population structure analysis, we computed pairwise genetic distances between relevant pair of samples. For each sample, a within-sample non-reference genotype frequency (*g*_*s*_) was assigned at each barcode position. If the sample carried the reference allele, 0 is assigned; if the sample carried the alternative allele, 1 is assigned; if both alleles were present, 0.5 is assigned. Distance between two samples at that position was then calculated using the equation: *d* = *g*_1_(1 − *g*_2_) + *g*_2_(1 − *g*_1_), where *g*_1_ and *g*_2_ is the genotype frequency of sample 1 and sample 2 respectively. The pairwise distance was calculated as the mean of *d* across all loci where neither of the two samples had a missing call.

### Identification of highly related clusters in Attapeu

Samples collected in Attapeu between January 2011 and April 2021 were assigned into clusters of three or more similar parasites. Clustering was based on their pairwise genetic barcode similarity *s*, obtained from the pairwise genetic distance *d* where s=(1-*d*). Using the igraph R package^38^ (https://igraph.org/r/), we constructed a graph connecting sample pairs with *s* greater than a minimum threshold *s*_*min*_, and applied the Louvain multilevel community detection algorithm^39^ to partition the graph into clusters. In the present analysis, we used *s*_*min*_=0.95 to obtain clusters of parasites with essentially identical barcodes, allowing for a low proportion of genotyping errors.

To assess whether the outbreak clusters were present outside of southern Laos, we also performed clustering after including publicly available GRC data from 9,609 samples collected from eight countries by the GenRe-Mekong Project^27^. Spatial distribution of highly-related clusters was visualized by producing prevalence maps using bespoke R code using map data from Natural Earth (https://www.naturalearthdata.com/).

### Characterization of population-level genetic diversity

Population-level genetic diversity was assessed by the mean expected heterozygosity 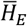 across all barcode SNPs. At each barcode SNP, the expected heterozygosity *H*_*E*_ (the proportion of heterozygous genotypes expected under Hardy-Weinberg equilibrium^40^) was estimated using the equation: 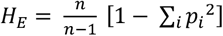, where *n* = the number of samples in the population that carried a valid genotype at this SNP, and *p*_*i*_ = the allele frequency of the *i*th allele observed. Missing alleles were removed before computing allele frequencies (*p*_*i*_) and heterozygous alleles contributed by 0.5 to each of the nucleotide allele in the locus.

### Statistical analysis

All statistical analyses were performed using R (V4.2.0). Statistical comparisons were conducted using Student’s *t* tests for continuous variables between two groups. One-way ANOVA and Tukey’s HSD post hoc test was used to compare *H*_*E*_ between populations. *P* values <0.05 were considered statistically significant.

To determine which *kelch13* genotypes were associated with the outbreak in Attapeu, covariates were entered into a logistic regression model. Linear regression was performed using glm function in R, with outbreak outcomes as the dependent variable and *kelch13* genotypes as the predictors. Exponentiating the linear regression coefficient provided the odds ratio, which gives a measure of the impact of changing a *kelch13* genotype on the odds of an outbreak occurring.

### Population structure analysis

To visualize relationships between samples, heatmaps of pairwise distance matrices were produced using the Heatmap function in the ComplexHeatmap R package (V2.8.0).^41^ The arrangement of samples on the heatmap was determined by hierarchical clustering using the shortest distance between two samples (clustering_distance=‘euclidean’) to maximize the distance between two clusters (clustering_method=‘complete’). Annotations were passed onto the heatmap to provide sample metadata including: collected year, *kelch13* mutation, *plasmepsin2/3* amplification status, and cluster label or country of collection. The same pairwise genetic distance matrix was used to perform principal coordinate analysis (PCoA) using the cmdscale function of the ape R package.^42^ Samples that did not cluster with any other sample were excluded in the PCoA.

### Ancestry analysis

Using genotypes obtained from WGS data for all PfCP samples from Laos, Cambodia, Vietnam and northeast Thailand, we identified 53,150 SNPs for which there was at least one sample with a homozygous call for the reference allele, and at least one with a homozygous call for an alternative allele. We used these genotypes to estimate alternative allele frequencies; missing genotypes were excluded in this estimation. Identity by descent (IBD) analysis was performed on the resulting genotypes and allele frequencies, using hmmIBD^43^ with default parameters. Visualizations of recombination patterns were produced from hmmIBD results using bespoke R scripts.

## Supporting information

Study Sites Credits

## Data Availability

All data produced in the present study are available upon request and will be made publicly available at malariagen.net upon publication in a peer reviewed journal.

## Acknowledgments

The authors wish to thank all the patients who generously agreed to provide blood samples, and their guardians. We also wish to thank all public health and medical staff who participated in the collection of samples used in this study; a full list of sites and local collaborators is included in the Supplementary Materials. Genome sequencing and genotyping was performed by the Wellcome Sanger Institute (WSI), and sequencing data processing was supported by the MalariaGEN Resource Centre. We thank the staff of the WSI Sample Logistics, Sequencing, and Informatics facilities for their contribution; Eleanor Drury for the support in the sample processing pipeline; Victoria Simpson and Kim Johnson for coordinating the MalariaGEN Resource Centre. This publication uses data from the MalariaGEN *Plasmodium falciparum* Community Project as described in ‘An open dataset of *Plasmodium falciparum* genome variation in 7,000 worldwide samples. MalariaGEN et al, Wellcome Open Research 2021, 6:42 (https://doi.org/10.12688/wellcomeopenres.16168.2)’. Maps were made with Natural Earth. Free vector and raster map data @ naturalearthdata.com.

