## Supplementary Materials for "Genetic analysis of a malaria outbreak in Laos driven by a selective sweep for *Plasmodium falciparum kelch13* R539T mutants"

### Supplementary Tables

| | | | Number of samples before filtering | | | | | | Number of samples after filtering | | | | | |
|---|---|---|---|---|---|---|---|---|---|---|---|---|---|---|
| **Source** | **Study** | **Year** | **Attapeu** | **Champasak** | **Salavan** | **Savannakhet** | **Sekong** | **Total** | **Attapeu** | **Champasak** | **Salavan** | **Savannakhet** | **Sekong** | **Total** |
| TRAC [6] | 1052-PF-TRAC-WHITE | 2011 | 59 | | | | | 59 | 59 | | | | | 59 |
| | | 2012 | 27 | | | | | 27 | 26 | | | | | 26 |
| GenRe-Mekong | 1208-PF-LA-CMPE-GENRE | 2017 | 97 | 94 | 102 | 272 | 13 | 578 | 86 | 85 | 100 | 259 | 12 | 542 |
| | | 2018 | 203 | 139 | 98 | 374 | 9 | 823 | 192 | 136 | 92 | 341 | 8 | 769 |
| | | 2019 | 48 | 28 | 27 | 209 | 11 | 323 | 39 | 21 | 15 | 121 | 7 | 203 |
| | | 2020 | 210 | 2 | 12 | 64 | 2 | 290 | 157 | 2 | 9 | 53 | 1 | 222 |
| | | 2021 | 64 | | | | | 64 | 51 | | | | | 51 |

**Supplementary Table 1. Number of samples collected in five provinces in Southern Laos.**

The table shows in the first two columns the project and study that contributed the samples; then for each year of collection, it shows the number of samples collected in each of the five provinces, and the total. Numbers are provided for the samples contributed (blue headers), and samples used in analyses after quality filtering (i.e. after removing samples with >25% genotype missingness in the genetic barcodes, yellow headings)

**(Next page)**

**Supplementary Table 2. Temporal distribution and genotype summary of 30 clusters identified in Attapeu.**

For each cluster, we show: the cluster label; the number of samples per year of collection; the total number of samples in the cluster; and the genotypes/haplotypes at key drug resistance-related loci: *kelch13* (resistance to artemisinin), *plasmepsin 2/3* (piperaquine), *pfcrt* (chloroquine), *pfdhfr* (pyrimethamine); *pfdhps* (sulfadoxine); and *pfmdr1* (amodiaquine). Full details about these haplotypes are given in the SpotMalaria Technical Notes at https://www.malariagen.net/resource/29.

| Cluster | 2011 | 2012 | 2017 | 2018 | 2019 | 2020 | 2021 | Total | Kelch13 | Pm2/3 | PfCRT | PfDHFR | PfDHPS | PfMDR1 |
|---|---|---|---|---|---|---|---|---|---|---|---|---|---|---|
| LAA1 | | | 3 | 9 | 5 | 108 | 43 | 168 | R539T:162 ; C580Y:1 | WT | CVIET | IRNI | AGEAA | NFD |
| LAA2 | | | 1 | 1 | 1 | 18 | 4 | 25 | C580Y:22 | WT | CVIET | IRNL | SGNGA | NFD |
| LAA3 | | | | 25 | | 1 | | 26 | WT: 26 | WT | CVIDT | IRNI | SAKAA | NYD |
| LAA4 | | | | 24 | 3 | | | 27 | WT: 27 | WT | CVIDT | IRNI | SGEAA | YYD |
| LAA5 | | | 4 | 18 | 2 | 2 | | 26 | C580Y:25 | Amplified | CVIET | IRNL | SGNGA | NFD |
| LAA6 | | | | 8 | 7 | 4 | | 19 | WT: 19 | WT | CVIDT | IRNI | SGKAA | NYD |
| LAA7 | | | | | 7 | 8 | 1 | 16 | R539T:16 | WT | CVIET | IRNL | SGNGA | NFD |
| LAA8 | | | 14 | 3 | | | | 17 | WT:6 ; C580Y:1 | WT | CVIDT | IRNI | SGEAA | YYD |
| LAA9 | | | | 8 | 1 | | | 9 | C580Y:9 | Amplified | CVIET | IRNI | SGNGA | NFD |
| LAA10 | | | | 10 | | | | 10 | C580Y:10 | WT | CVIDT | IRNI | AGEAT | NFD |
| LAA11 | | | 4 | | 3 | | | 7 | C580Y:5 | Amplified | CVIET | IRNL | SGNGA | NFD |
| LAA12 | | | 7 | | | | | 7 | C580Y:1 | WT | CVIDT | IRNI | AGEAT | NYD |
| LAA13 | | | 7 | | | | | 7 | C580Y:5 | Amplified | CVIET | IRNL | AGEAA | NFD |
| LAA14 | | | | 2 | 3 | | | 5 | C580Y:5 | Amplified | CVIET | IRNL | SGNGA | NFD |
| LAA15 | 2 | 3 | | | | | | 5 | WT:5 | WT | CVIET | NRNI | AGEAA | NYD |
| LAA16 | | | | | | 4 | | 4 | C580Y:4 | WT | CVIET | IRNI | SGKAA | NFD |
| LAA17 | | | | 4 | | | | 4 | WT:4 | WT | CVIDT | IRNI | SAKAA | NYD |
| LAA18 | | | | 3 | | | | 3 | C580Y:3 | Amplified | CVIET | IRNL | SGNGA | NFD |
| LAA19 | | | | 1 | | 2 | | 3 | C580Y:3 | WT | CVIET | IRNI | SGKAA | NFD |
| LAA20 | | | 4 | | | | | 4 | WT:3 | WT | CVIDT | IRNI | FAKAS | NYD |
| LAA21 | | | | | | 1 | 2 | 3 | WT:3 | WT | CVIET | IRNI | FAKAS | NFD |
| LAA22 | | | | 1 | | 2 | | 3 | WT:3 | WT | CVIET | IRNI | FAKAS | NYD |
| LAA23 | | | | 3 | | | | 3 | WT:3 | WT | CVIDT | IRNI | SGKAA | NYD |
| LAA24 | | | | 3 | | | | 3 | C580Y:3 | Amplified | CVIDT | IRNL | AGEAT | NFD |
| LAA25 | | | | 3 | | | | 3 | WT:3 | WT | CVIET | IRNI | *G*A* | NYD |
| LAA26 | | | | 3 | | | | 3 | C580Y:3 | Amplified | CVIET | IRNL | SGNGA | NFD |
| LAA27 | | | | 3 | | | | 3 | C580Y:3 | Amplified | CVIET | IRNI | AGEAA | NYD |
| LAA28 | | | 2 | 1 | | | | 3 | WT:2 | WT | CVIDT | IRNI | AGEAT | NYD |
| LAA29 | | | 3 | | | | | 3 | WT:3 | WT | CVIDT | IRNI | AGKAT | NYD |
| LAA30 | | | 3 | | | | | 3 | | WT | CVIET | IRNI | SAKAA | NYD |
| Samples in cluster | 2 | 3 | 52 | 133 | 32 | 150 | 50 | 422 | | | | | | |
| Unique samples | 57 | 23 | 34 | 59 | 7 | 7 | 1 | 188 | | | | | | |
| *Total* | *59* | *26* | *86* | *192* | *39* | *157* | *51* | ***610*** | | | | | | |

| Covariate | Odds ratio | z | p Value | 95% CI |
|---|---|---|---|---|
| R539T | 52.8 | 7.055 | <0.0001*** | 19.5-186.0 |
| Het | 6.67 | 1.802 | 0.072 | 0.72-54.5 |
| C580Y | 1.8 | 1.037 | 0.300 | 0.65-6.41 |
| WT | 0.33 | -1.726 | 0.084 | 0.094-1.29 |

**Supplementary Table 3. Multivariate logistic regression of kelch13 genotypes associated with Attapeu outbreak.**

For each *kelch13* allele observed in the Attapeu population, we report the odds ratio that it is associated with the outbreak. Other statistics (z-value, p-value and 95% confidence interval) are also reported.

| Label | Kelch13 | Amplifications | Province, Country | Year | Description | PfCP Release | MalariaGEN Id | ENA Id |
|---|---|---|---|---|---|---|---|---|
| **LAA1** | R539T | m+ p- | Attapeu, Laos | 2018 | Attapeu outbreak strain LAA1 | 6.3 | RCN13530 | ERS2866227 |
| **LAA2** | C580Y | m- p- | Attapeu, Laos | 2018 | Attapeu outbreak strain LAA2 | 6.3 | RCN13540 | ERS2866240 |
| **LAA7** | R539T | m- p- | Attapeu, Laos | 2019 | Attapeu outbreak strain LAA7 | 6.4 | RCN25946 | ERR5481228 |
| **KH3** | R539T | m+ p- | Battambang, Cambodia | 2008 | Early ART-R, KH3 group | 6.0 | PH0145-CW | ERS024144 |
| **KH2A** | C580Y | m- p- | Battambang, Cambodia | 2009 | Early ART-R, KH2 group | 6.0 | PH0147-CW | ERS024146 |
| **KH2B** | C580Y | m- p+ | Pursat, Cambodia | 2010 | Early ART-R, KH2 group (KEL1/PLA1 precursor) | 6.0 | PH0169-C | ERS014171 |
| **KH3-LA** | R539T | m+ p- | Champasak, Laos | 2017 | Likely major contributor to LAA1 | 6.2 | RCN08982 | ERS2474155 |
| **LAA1-pre** | R539T | m+ p- | Attapeu, Laos | 2018 | Likely LAA1 precursor, with *mdr1* amplification | 6.3 | RCN13530 | ERS2866227 |
| **LAA1-preWT** | WT | m- p- | Champasak, Laos | 2018 | Likely contributor to LAA1 (Laos WT parasite) | 6.3 | RCN13467 | ERS2866164 |
| **LAA2-pre** | C580Y | m? p- | Champasak, Laos | 2018 | Identical to LAA2, found in Laos before outbreak | 6.4 | RCN15245 | ERR3831106 |
| **LAA7-preA** | R539T | m- p- | Attapeu, Laos | 2018 | Likely contributor to LAA7 (R539T) | 6.3 | RCN11913 | ERS2463794 |
| **LAA7-preB** | C580Y | m- p+ | Attapeu, Laos | 2018 | Likely contributor to LAA7 (C580Y) | 6.3 | RCN13527 | ERS2866224 |

**Supplementary Table 4. Metadata for selected samples used to reconstruct the ancestry of the Attapeu outbreak strains.**

Whole-genome sequencing data for these samples was used to identify IBD proportions and patterns. For each sample, we show: the label used to refer to the sample in the main text and in Figure 4; the *kelch13* allele; the amplification status for the *pfmdr1* (m) and *plasmepsin 2/3* (p) loci (a + sign indicates amplification, a - indicates single copy, a ? denotes an undetermined result); the province and country of collection; the year of collection; a description of the sample's significance in the ancestry analysis; the *P. falciparum* Community Project release that included the sample; its MalariaGEN identifier; and the sample's identifier at the European Nucleotide Archive (ENA) where the sequencing data is deposited.

### Supplementary Figures

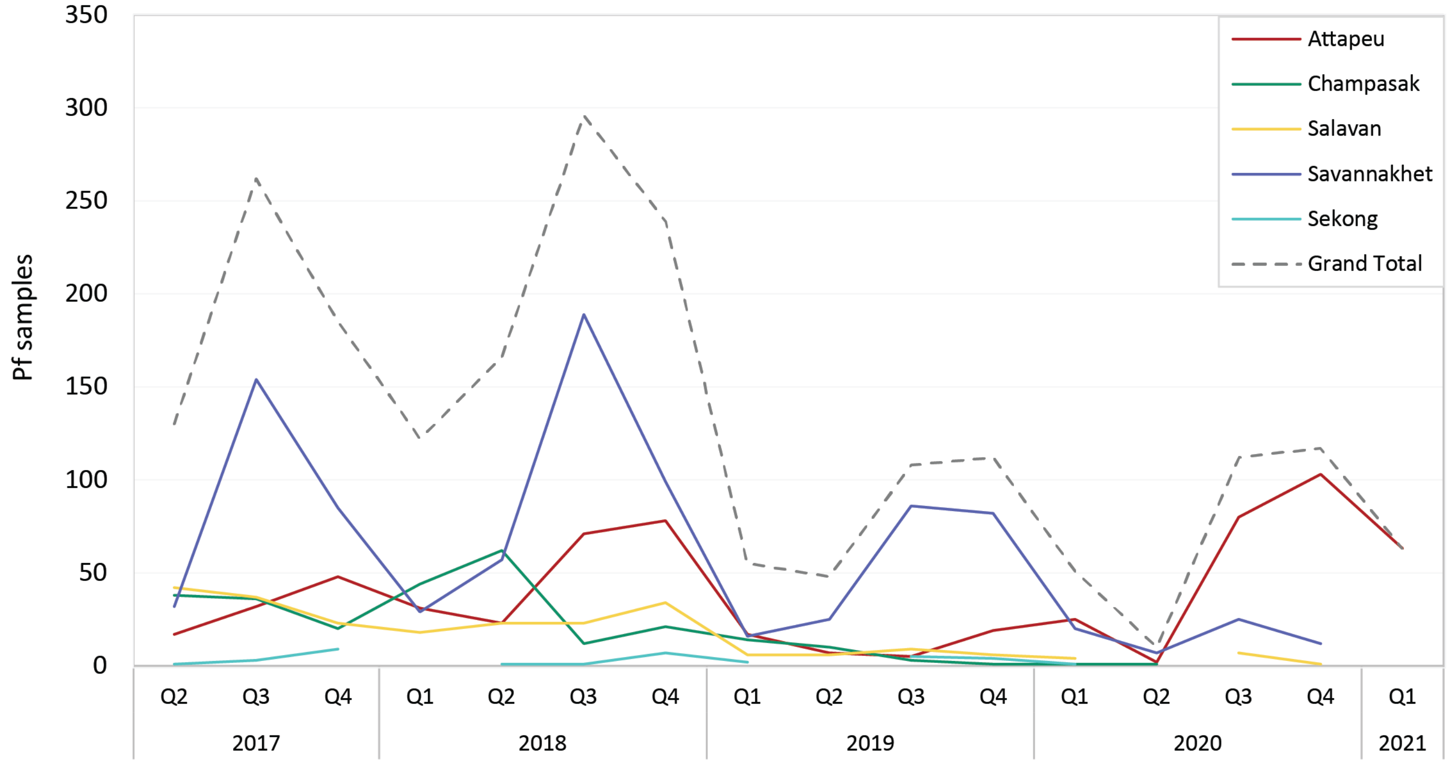

**Supplementary Figure 1. Number of *Plasmodium falciparum* samples in Laos collected by GenRe-Mekong (Apr 2017 - Mar 2021).**

The graph shows the number of samples collected in each quarter in each of the five endemic provinces of Laos. The dashed line shows the total number of samples collected. Q1: January – March; Q2: April – June; Q3: July – September; Q4: October –December.

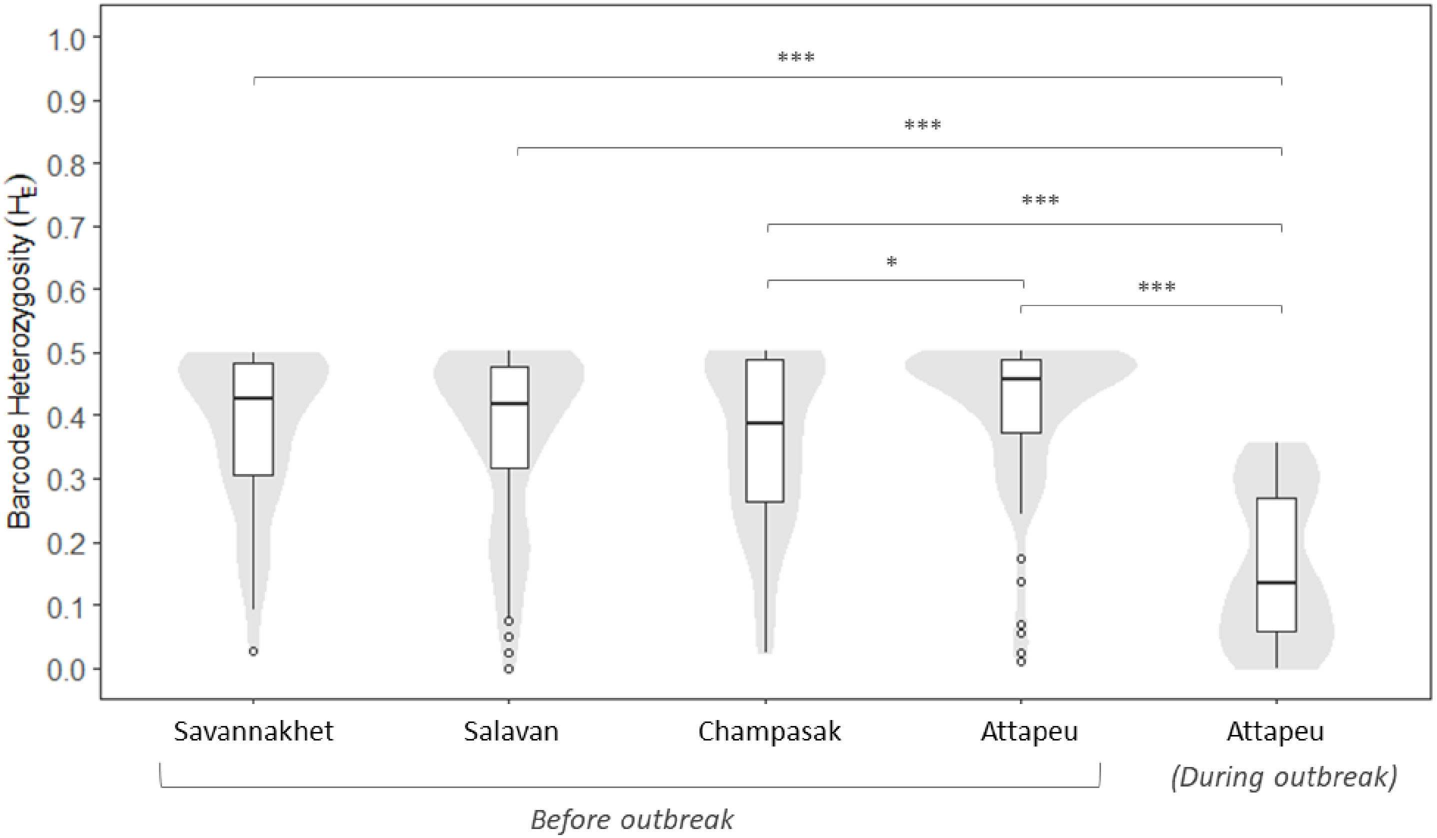

**Supplementary Figure 2. Barcode heterozygosity before and during the outbreak across four provinces in Laos.**

Density plots of the value distributions is shown in grey behind the boxplots. Mean comparisons by Tukey's HSD post-hoc test between groups are indicated by horizontal lines and asterisks. Significance codes: *** p < 0.001*, **p < 0.01, *p < 0.05*. A highly significant reduction in barcode heterozygosity is observed in Attapeu province during the outbreak.

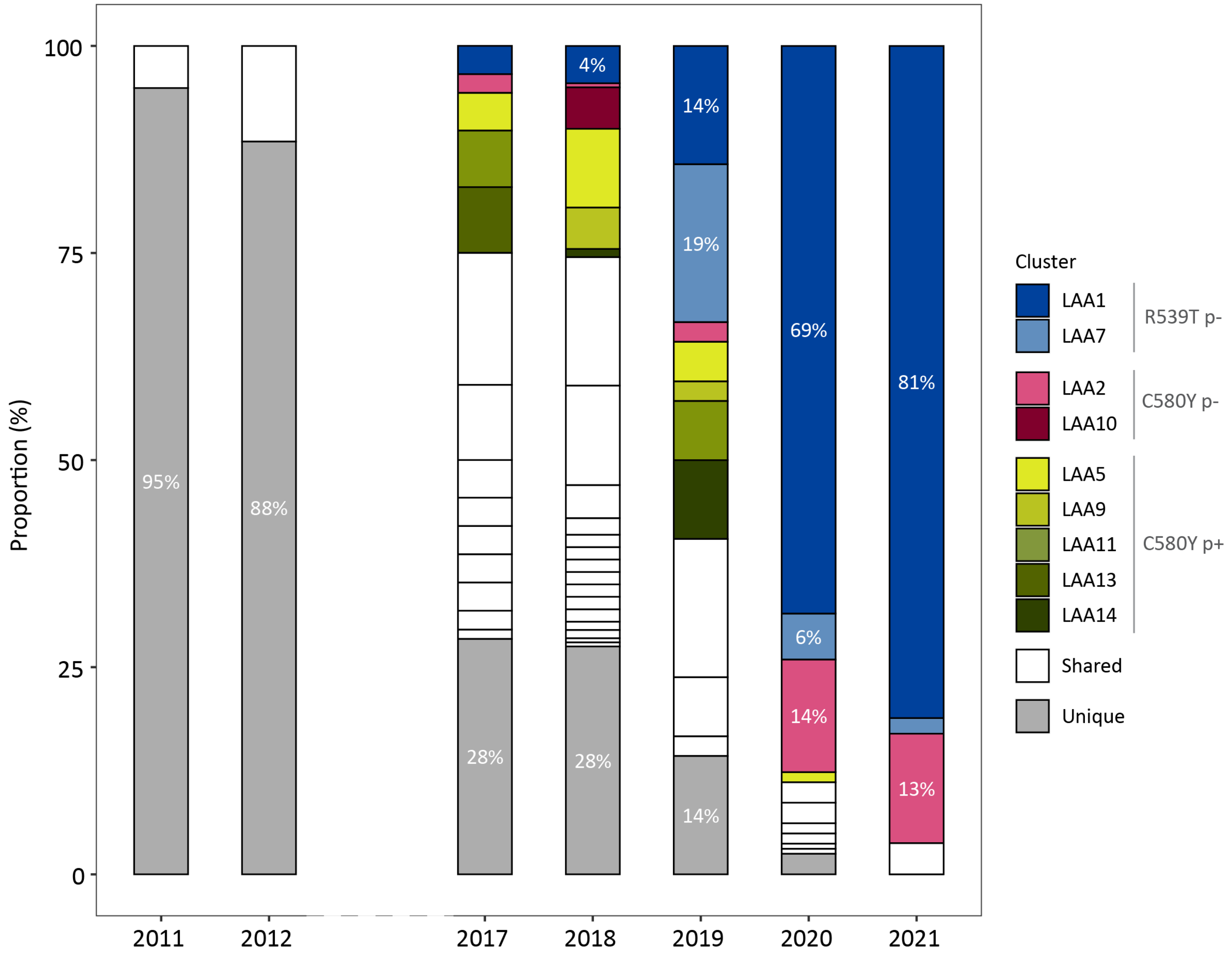

**Supplementary Figure 3. Prevalence of *kelch13* clusters in Attapeu**

Clusters contain samples that share at least 95% barcode identity (see Methods). Colours are given to clusters of at least 5 members in which at least 5 members carry a *kelch13* mutation (R539T or C580Y). Percentage is shown only for LAA1, LAA2 and LAA7 groups when ≥ 4%. As shown in the legend, clusters are grouped according to their *kelch13* genotype and by their *plasmepsin2/3* amplification status (p- and p+ denoting single- and multiple-copy samples), and coloured accordingly. Other clusters (containing *kelch13* wild-type parasites) are not coloured, and labelled as Shared. Parasites that did not form part of any cluster are grouped together in a gray segment, and labelled as Unique.

**(Next Page)**

**Supplementary Figure 4. Prevalence of the outbreak populations in the Greater Mekong subregion.**

Prevalence and cluster statistic for clusters (A) LAA1, (B) LAA2, and (C) LAA7. Circular markers show the prevalence among all samples from the same province. Provinces are connected by lines whose thickness provides a qualitative visualization of the level of sharing. For LAA1 and LAA7, which are limited to Attapeu province, smaller maps on the right of the panel show the prevalence by district. A large proportion of the population in Attapeu (36%) belongs to the LAA1 cluster, with Phouvong district as the hotspot of transmission. In contrast, LAA2 parasites have been circulating across several GMS countries, albeit at very low frequency.

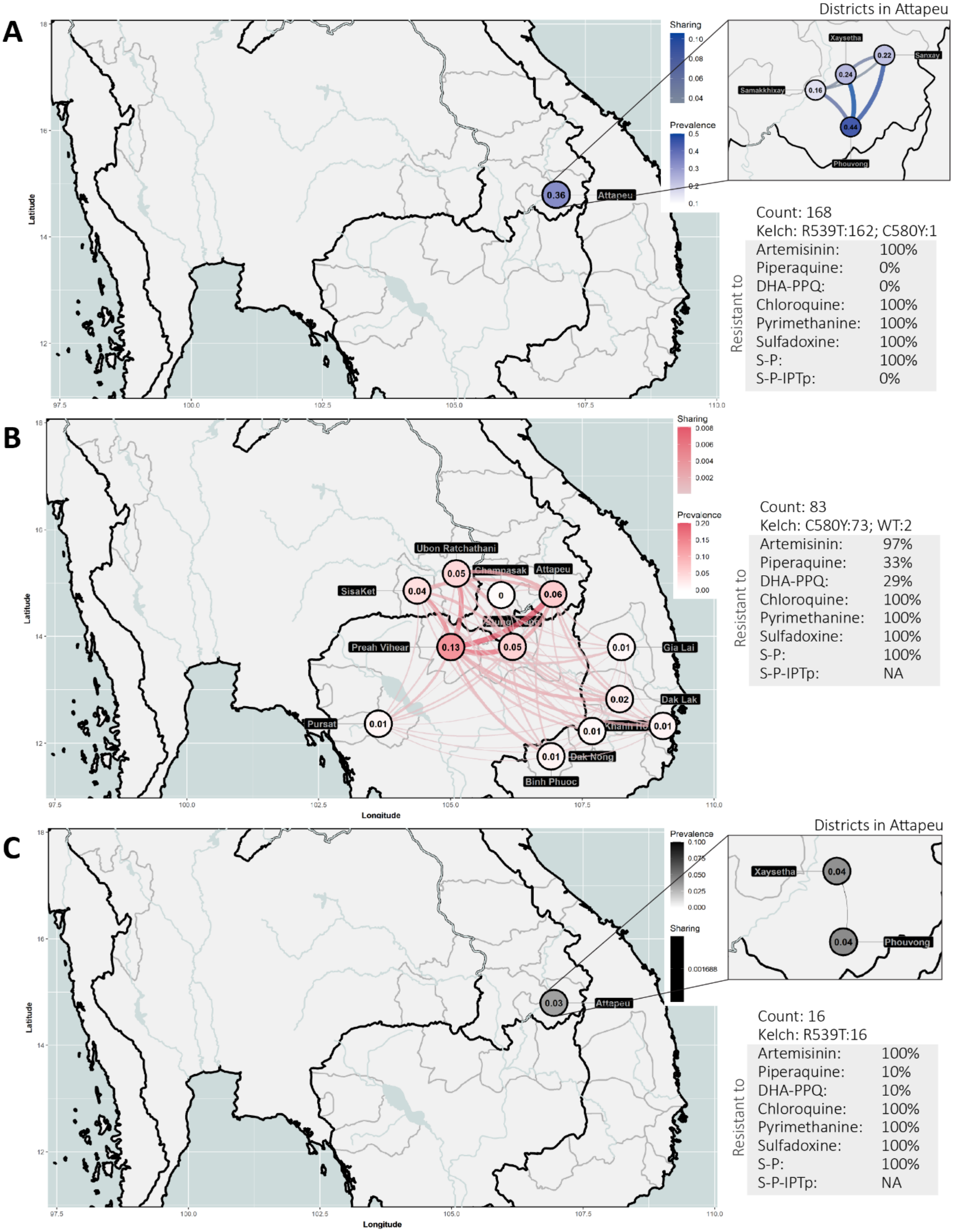
A
Districts in Attapeu
Sharing
Prevalence
Latitude
Attapeu
0.36
Xaysetha
Sanxay
Samakkhixay
Phouvong
0.22
0.24
0.16
0.44
Count: 168
Kelch: R539T:162; C580Y:1
Resistant to
Artemisinin: 100%
Piperaquine: 0%
DHA-PPQ: 0%
Chloroquine: 100%
Pyrimethanine: 100%
Sulfadoxine: 100%
S-P: 100%
S-P-IPTp: 0%
B
Sharing
Prevalence
Latitude
Longitude
Ubon Ratchathani
Champasak
Attapeu
SisaKet
Preah Vihear
Gia Lai
Dak Lak
Pursat
Dak Nong
Binh Phuoc
0.05
0.04
0
0.06
0.13
0.05
0.01
0.02
0.01
0.01
0.01
0.01
Count: 83
Kelch: C580Y:73; WT:2
Resistant to
Artemisinin: 97%
Piperaquine: 33%
DHA-PPQ: 29%
Chloroquine: 100%
Pyrimethanine: 100%
Sulfadoxine: 100%
S-P: 100%
S-P-IPTp: NA
C
Districts in Attapeu
Prevalence
Sharing
0.001688
Latitude
Longitude
Attapeu
0.03
Xaysetha
Phouvong
0.04
0.04
Count: 16
Kelch: R539T:16
Resistant to
Artemisinin: 100%
Piperaquine: 10%
DHA-PPQ: 10%
Chloroquine: 100%
Pyrimethanine: 100%
Sulfadoxine: 100%
S-P: 100%
S-P-IPTp: NA

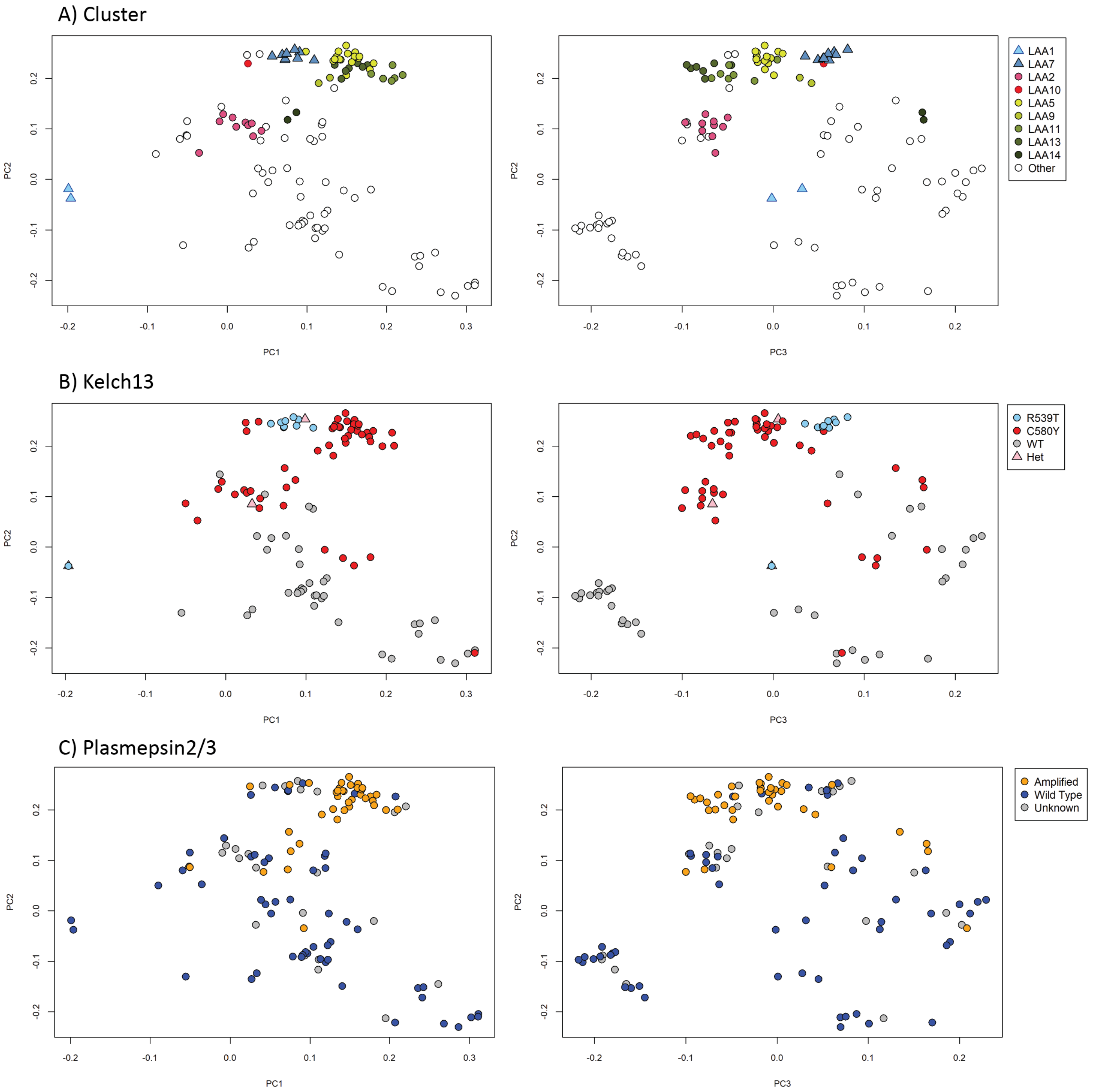

**Supplementary Figure 5. Population structure of Attapeu populations shown on PCoA**

In this PCoA analysis, samples are colored according to (A) their assigned cluster, (B) *kelch13* mutation, and (C) *plasmepsin 2/3* amplifications. Only samples belonging to clusters were included in this analysis. The first three principal coordinates explained 32.2%, 20.6% and 9.5% of the variance, respectively. LAA1 shows clear separation from other clusters and a high degree of identity. Although LAA2 carries the C580Y *kelch13* mutation, it groups separately from earlier KEL1/PLA1 parasites, while LAA7 is more closely related in spite of its R539T mutation.

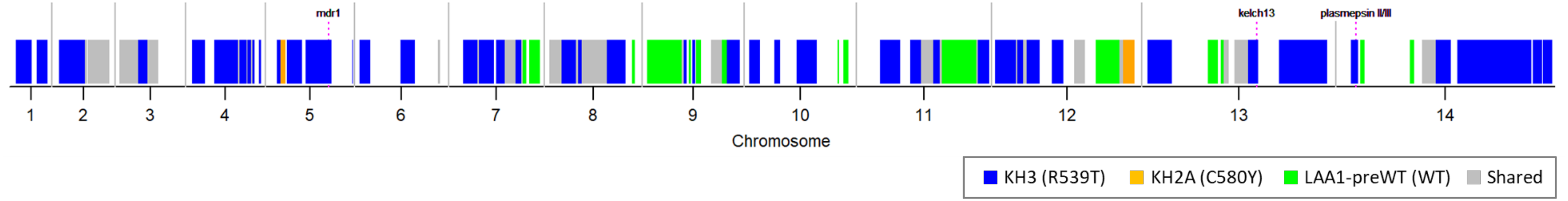

**Supplementary Figure 6. Contributions to the LAA1 genome by three different genetic sources.**

The plot shows the 14 nuclear chromosomes of P. falciparum along the horizontal axis, their size proportional to the number of bases. The chromosomes are separated by gray vertical lines, and dotted lines show the location of the *kelch13* gene and of the *plasmepsin 2/3* and *mdr1* amplifications. Blocks of the LAA1 genome are coloured according to whether they are in IBD with KH3 (blue), KH2A (orange), LAA1-preWT (green) or more than one of these (gray). Regions without a colour block are not in IBD with any of the three strains tested.

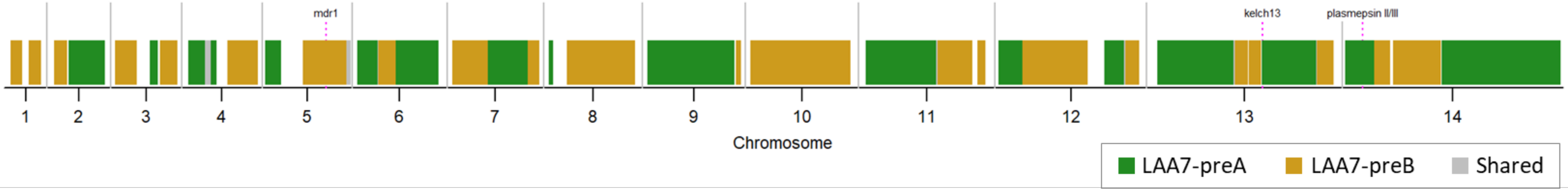

**Supplementary Figure 7. Contributions to the LAA7 genome by two pre-outbreak strains.**

The plot shows the 14 nuclear chromosomes of P. falciparum along the horizontal axis, their size proportional to the number of bases. The chromosomes are separated by gray vertical lines, and dotted lines show the location of the *kelch13* gene and of the *plasmepsin 2/3* and *mdr1* amplifications. Blocks of the LAA7 genome are coloured according to whether they are in IBD with LAA7-preA (R539T, green), LAA7-preB (C580Y, brown) or both (gray). Regions without a colour block are not in IBD with either strain.
