## Supplementary material for "Genetic analysis of a malaria outbreak in Laos driven by a selective sweep for *Plasmodium falciparum kelch13* R539T mutants": Study Sites Credits

### Study sites and local collaborators in the Lao PDR

We thank the following individuals at the study sites in the Lao PDR for their input to the present outbreak study as well as for their contributions to the GenRe-Mekong Project.

#### Attapeu Province

##### Collaborators

| | |
|---|---|
| Mr. Sengdala Phetsomphou | Mr.Philavong Dethphachan |
| Mrs. Khamlar Khounxay | Mr. Bounkan Vongsamlan |
| Mr. Phou Ngeun Xaiyavong | Mr. Kethsana Xaiyasaeng |
| Mr. Boudmanda Vongphanu | Mr. Simsa Nguan Saenthuliboud |
| Ms. Ari Phommachan | Mrs. Nuthong Sibounhueng |
| Mrs. Nali Xaiyaserm | Ms. Oudomhak Singsombus |
| Mr. Phouvieng Xinalath | Mr. Kawin Xaybounmee |

##### Study sites

LA500 Attapeu PH, Attapeu, Attapeu
LA502 Attapeu Provincial Military Hospital, Attapeu, Attapeu
LA510 Xaysetha DH, Xaysetha, Attapeu
LA511 Keng Yai HC, Xaysetha, Attapeu
LA512 Phon Gnam HC, Xaysetha, Attapeu
LA513 KhengMak-Keun HC, Xaysetha, Attapeu
LA514 PhaoSamPhanMeeXay HC, Xaysetha, Attapeu
LA515 SaPhouan HC, Xaysetha, Attapeu
LA520 Sanamxay DH, Sanamxay, Attapeu
LA521 Somsouk HC, Sanamxay, Attapeu
LA522 Sompoy HC, Sanamxay, Attapeu
LA530 Xanxay DH, Xanxay, Attapeu
LA540 Phouvong DH, Phouvong, Attapeu
LA541 Phouhome HC, Phouvong, Attapeu

#### Champasak Province

##### Collaborators

| | |
|---|---|
| Mr. Saitadam Phetxilar | Mrs. Kularb Keokounmuong |
| Mrs. Aran Souniyavong | Mr. Sam Keomanivong |
| Mrs. Phetsamon Sisavath | Mrs. Amphon Keomanee |
| Mrs. Kanhmany Vongphoumee | Mr. Phoukhanh Chanthavisin |
| Mrs. Phaitoun Inthathilath | Mrs. Kaikham Inthathirath |
| Mr. Toulee Saivaya | Mr. Chanthala Sisomvang |
| Mrs. Vilayvanh Vongphoumee. | Mr.Thongbai Phommixay |
| Mr. Khamphong Luangphasee | Mrs. Soutthanong Sisamoud |
| Mr. Amphon Vongsounthone | Mrs. Vanthong Somchidkhamluxay |
| Mrs. Soviet Pasidson | Mrs. Navalit Sivilay |
| Mr. Shai Namxokchid | |

##### Study sites

| | |
|---|---|
| LA300 Pakse PH, Pakse, Champasak | LA331 Nong nga HC, Mounlapamok, Champasak |
| LA303 Champasak Provincial 106 Technical of Military Hospital, Pakse, Champasak | LA332 Nadee HC, Mounlapamok, Champasak |
| LA310 Paksong DH, Paksong, Champasak | LA340 Khong DH, Khong, Champasak |
| LA311 Pakpong HC, Paksong, Champasak | LA341 Nafang HC, Khong, Champasak |
| LA312 Huaykong HC, Paksong, Champasak | LA342 Phonsa arth HC, Khong, Champasak |
| LA320 Pathoumphone DH, Pathoumphone, Champasak | LA343 Ban Soth HC, Khong, Champasak |
| LA321 Phapho HC, Pathoumphone, Champasak | LA350 SaNaSomBoun DH, SaNaSomBoun, Champasak |
| LA322 Sanod HC, Pathoumphone, Champasak | LA360 ChamPaSak DH, ChamPaSak, Champasak |
| LA323 Palai HC, Pathoumphone, Champasak | LA361 NongTae HC, ChamPaSak, Champasak |
| LA330 Mounlapamok DH, Mounlapamok, Champasak | LA370 SouKhoumMa DH, SouKhoumMa, Champasak |

### Salavan Province

#### Collaborators

| | |
|---|---|
| Mr. Keomany | Mr. Lamphanh Salapheung |
| Mrs. Kaikeo Duongxilee | Mr. Bounlom Phonsaly |
| Mrs. Keochai | Mr. Khammanh Phommixay |
| Ms. Khamkak Xaiyason | Mr. Somchit Keochomlath |
| Mrs. Vilayvong Sengkeonee | Mrs. SomThid Molixath |
| Mrs. Eatsaphone Duongphasouk | Mrs. Kanha Souliya |
| Mr. Mad Khamsouk | Mrs. Singha |
| Ms. Ximuon Xixana | Mr. Kuta Keotokong |
| Mrs. Chanthaly Soukhamthud | Mr. Boualapha Khamcounmuong |
| Mr. Phetsamai Getsombus | Mr. Savanthong Keoduongsee |

#### Study sites

LA200 Salavan PH, Salavan, Salavan

LA202 Salavan Provincial Military Hospital, Salavan, Salavan

LA210 Ta Oy DH, Ta Oy, Salavan

LA211 Phoutang HC, Ta Oy, Salavan

LA212 Pajudone HC, Ta Oy, Salavan

LA213 SoyTamh HC, Ta Oy, Salavan

LA214 KokBok HC, Ta Oy, Salavan

LA220 Toomlarn DH, Toomlarn, Salavan

LA221 Tabeng HC, Toomlarn, Salavan

LA230 Vapy DH, Vapy, Salavan

LA231 Khonsai HC, Vapy, Salavan

LA232 Saphab HC, Vapy, Salavan

LA240 Samuoi DH, Samuoi, Salavan

LA241 Asok HC, Samuoi, Salavan

LA242 Axing HC, Samuoi, Salavan

LA243 KiNae HC, Samuoi, Salavan

LA244 AhWao HC, Samuoi, Salavan

LA245 Khongsedone DH, Khongsedone, Salavan

LA246 Lakhonepheng DH, Lakhonepheng, Salavan

### Sekong Province

#### Collaborators

| | |
|---|---|
| Mrs. Sonsamai Xaiyavong | Mr. Siamphai Canlamphan |
| Mrs. Thidthisouk Kongsavanh | Mr. Huck Kampouxa |
| Mr. Phoudsy Xaysueksa | Mrs. Jhonlakham Soulivanh |
| Mrs. Sengkham Phonvilay | Mr. SantiPhone Xaiyason |
| Mrs. Daovaly Dalavong | Mr. Kamxi Vongvaengouk |
| Mr. Phou Ngeun Doungmala | Mr. SanDey Leuymalaysee |
| Mrs. Orlaphan Souvannasee | Mrs. Sidda Amphaiphon |
| Mrs. Vanthana Inthamixay | Mr. Pethsamai |

#### Study sites

LA400 Sekong PH, Sekong, Sekong

LA402 Sekong Provincial Military Hospital, Lamam, Sekong

LA410 Thateng DH, Thateng, Sekong

LA411 Thonnoy HC, Thateng, Sekong

LA420 Kaleum DH, Kaleum, Sekong

LA421 Jii HC, Kaleum, Sekong

LA430 Dakcheung DH, Dakcheung, Sekong

LA431 Dakdun HC, Dakcheung, Sekong

LA442 Ban Phone HC, Lamam, Sekong

LA443 NaKhasangkang HC, Lamam, Sekong

LA444 TaNeup HC, Lamam, Sekong

LA445 Tock Ongkeo HC, Lamam, Sekong

LA446 DoneChanh HC, Lamam, Sekong

LA447 Ban Ngeup HC, Lamam, Sekong

#### Savannakhet Province

##### Collaborators

| | |
|---|---|
| Mrs. Vilayphone Pha Arnan | Mrs. Phansida chanthilanong |
| Mrs. Phaivanh Sykhammounty | Mrs. Nilaphon OunLeuyvongsak |
| Mrs. Vanna Saengchan | Mrs. Nui Lianvilay |
| Mrs. Souksavanh | Mrs. Malina Xaiyavong |
| Mrs. Outhan Xiphommalangkoun | Ms. Sompharn Vannavong |
| Mr. Xisouphan Luangphasee | Ms. Phoukham Malavanh |
| Mrs. Khammany Saenboudpalath | Mr. Kongkeo Thammalin |
| Ms. Rodjana Boudsisavanh | Mr. Souksamon Thinlanong |
| Ms. Phouthon Souliyavong | Mrs. Sengsavanh Harnxana |
| Ms. Phoukhao Phudthavadee | Mrs. Leevon Xaiyacom |
| Mr. Chai Khamphiu | Mr. Bounhom Chanthaluxay |
| Ms. Authai Lukhamharn | Mr. Norchan |
| Ms. Kayson Sisaketkhammuon | Mr. Lounnee Xaiyavong |
| Mrs. Toukta Khankeo | Ms. Vivanxay Silathom |
| Mr. Sonphet Sidala | Ms. Immalar Lathxavongphon |
| Ms. Intong Saengsoulichan | Mr. Khamkai Phetluangsee |
| Mr. Vongphachan Bounmee | Mr. Bualom Bangmano |
| Ms. Souvanna Ludthaphon | Mrs. Ketsoudar Duongsavanh |

##### Study sites

LA100 Savannakhet PH, Savannakhet, Savannakhet

LA102 Savannakhet Provincial Military Hospital, Kaysone, Savannakhet

LA110 Phine DH, Phine, Savannakhet

LA111 Nathong HC, Phine, Savannakhet

LA112 Maiphousy HC, Phine, Savannakhet

LA113 Tang-Alai HC, Phine, Savannakhet

LA114 TounKamh HC, Phine, Savannakhet

LA115 HouyHoi HC, Phine, Savannakhet

LA120 Sepone DH, Sepone, Savannakhet

LA121 Phohai HC, Sepone, Savannakhet

LA122 Phabang HC, Sepone, Savannakhet

LA123 Dansavan HC, Sepone, Savannakhet

LA124 DongSaVanh HC, Sepone, Savannakhet

LA125 ManhChee HC, Sepone, Savannakhet

LA127 KhaToub HC, Sepone, Savannakhet

LA133 TangAhLai HC, Nong, Savannakhet

LA134 LaKhai HC, Nong, Savannakhet

LA135 KhaySone HC, Nong, Savannakhet

LA136 PhounMarkMee HC, Nong, Savannakhet

LA137 AhSing HC, Nong, Savannakhet

LA138 DaenViLay HC, Nong, Savannakhet

LA140 Thapanthong DH, Thapanthong, Savannakhet

LA141 Sepong HC, Thapanthong, Savannakhet

LA142 Xekeu HC, Thapanthong, Savannakhet

LA143 Phoumaly HC, Thapangthong, Savannakhet

LA144 Khathongneun HC, Thapangthong, Savannakhet

LA150 Vilabuly DH, Vilabuly, Savannakhet

LA151 Nammahi HC, Vilabuly, Savannakhet

LA160 Thaphalanxay DH, Phalanxay, Savannakhet

LA170 Nachantai HC, Phalanxay, Savannakhet

LA130 Nong DH, Nong, Savannakhet

LA131 Nakong HC, Nong, Savannakhet

LA132 Dongnasan HC, Nong, Savannakhet

LA171 Nakapong HC, Phalanxay, Savannakhet

LA172 Songkhone DH, Songkhone, Savannakhet

LA190 Xonboury DH, Xonboury , Savannakhet
